# Consumer-grade wearables and machine learning sensitively capture disease progression in amyotrophic lateral sclerosis

**DOI:** 10.1101/2023.03.28.23287869

**Authors:** Anoopum S. Gupta, Siddharth Patel, Alan Premasiri, Fernando Vieira

## Abstract

ALS causes degeneration of motor neurons, resulting in progressive muscle weakness and impairment in fine motor, gross motor, bulbar, and respiratory function. Promising drug development efforts have accelerated in ALS, but are constrained by a lack of objective, sensitive, and accessible outcome measures. Here we investigate the use of consumer-grade wearable sensors, worn on four limbs at home during natural behavior, to quantify motor function and disease progression in 376 individuals with ALS over a several year period. We utilized an analysis approach that automatically detects and characterizes submovements from passively collected accelerometer data and produces a machine-learned severity score for each limb that is independent of clinical ratings. The approach produced interpretable and highly reliable scores that progressed faster than the gold standard ALS Functional Rating Scale-Revised (−0.70 SD/year versus -0.48 SD/year), supporting its use as a sensitive, ecologically valid, and scalable measure for ALS trials and clinical care.

## Introduction

Novel therapeutic modalities are now aimed at proximal disease mechanisms in amyotrophic lateral sclerosis (ALS) and other neurodegenerative diseases^1,2^. One major barrier to the successful and efficient development of disease-modifying therapies for neurodegenerative disorders is a lack of objective clinical outcome measures that account for disease heterogeneity and can sensitively quantify disease progression over the duration of a clinical trial^3–5^. The standard tool for assessing disease severity in ALS clinical trials and clinical care is a semi-quantitative rating scale (ALS Functional Rating Scale-Revised^6,7^ or ALSFRS-R) that uses multiple choice questions to evaluate several behavioral functions (e.g., walking, handwriting, speech, swallowing). The assessment is most often completed by clinicians specializing in ALS^7,8^, however recent studies have shown high correlation between clinician-performed ALSFRS-R and at-home, patient-performed ALSFRS-R^9^. Clinician or patient-performed ALSFRS-R is a useful assessment of global motor function, however it is subjective, categorical, and is only performed intermittently over time, which limits its sensitivity for measuring disease change and contributes to the need for relatively large and expensive trials^10,11^. This is a particular challenge in rare disease and results in pressure to include relatively homogenous cohorts with faster rates of disease progression, which restricts participation of some individuals and may not be representative of the entire ALS population^12^.

There is a great opportunity to reduce the size and cost of ALS trials, increase the population of individuals who can participate, and accelerate the evaluation of promising therapeutics through the development of new categories of sensitive quantitative motor outcome measures^13–15^. Quantitative motor outcome measures may be task-based (i.e., measuring behavior during performance of a specific task) or task-free, where an individual’s natural behavior is measured passively and continuously at home. There has been recent development of several task-based approaches to quantify speech and limb function in ALS using scalable technologies at home^9,16–18^ and only a single report of a task-free approach in ALS using a waist-worn accelerometer^19^. Task-based measures, however, have some of the same limitations as rating scales in that they are based on a relatively small number of data samples and cannot easily account for diurnal and day-to-day variability, they rely on the participant’s ability and motivation to perform the task, and they are susceptible to learning and placebo effects.

Task-free assessment approaches which passively and continuously measure natural behavior at home using consumer grade devices have the potential to overcome these limitations and be transformative by making reliable and sensitive measures available at scale. Furthermore, they have the potential to produce measures that more closely reflect the day-to-day function of the individual by measuring the individual’s own selection of behaviors. However, the information obtained by the tool must be interpretable and meaningful to support its use in clinical trials or clinical care.

Here we demonstrate that a submovement-focused analysis of triaxial accelerometer data^20,21^, recorded from wrist and ankle sensors worn by hundreds of individuals with ALS at home during natural behavior, produces interpretable and robust measures of motor function and disease progression. We develop a machine learning approach to train a model that is sensitive to disease change by utilizing the information for how individuals’ sensor-based movement patterns change over time, rather than being constrained by existing clinical assessments such as ALSFRS-R. We show that the model’s severity estimates and longitudinal trajectories are reliable and consistent with ALSFRS-R, but are more sensitive than the clinical scale for measuring change over time. Thus, we demonstrate that objective, sensitive, and scalable measures of motor function and disease change can be obtained from passive analysis of everyday behavior using inexpensive wearable sensors.

## Results

### Overview of the dataset

We analyzed accelerometer data from wrist and ankle-worn sensors collected as part of the Precision Medicine Program launched by the ALS Therapy Development Institute (ALS-TDI) in 2014 (see Methods). Individuals were asked to wear a sensor on each wrist and ankle as much as possible for one week each month. Participants also performed a sequence of 5 limb-based exercises on alternating days, lasting a total of approximately 5 minutes. An analysis of accelerometer data collected only during these brief task-based assessments was previously reported^17^. Here, we analyze the entirety of accelerometer data collected at home as individuals performed their typical daily routine without any constraints. Participant data are shown in Figure 1A and dataset filtering steps are described in Figure 1B. Cross sectional analysis included 4637 sessions from 402 unique participants (376 ALS, 26 controls) with at least 24 hours of recorded accelerometer data from all four limbs. Longitudinal analysis was conducted using data from participants with at least three data collection sessions spanning a minimum of 0.75 years (188 ALS and 6 control participants). Submovement, activity bout, activity index, and spectral movement features (85 total) were extracted from each session as previously described^20,21^ (Figure 1C, Supplementary Table 1). Single feature analysis was performed on a subset of 24 key submovement (SM) features of interest. These included SM distance, peak velocity, and peak acceleration (8 features each). Mean and standard deviation were computed for short duration and long duration SMs in the primary and secondary directions of planar movement resulting in 8 features for each measurement type.

**Table 1.**
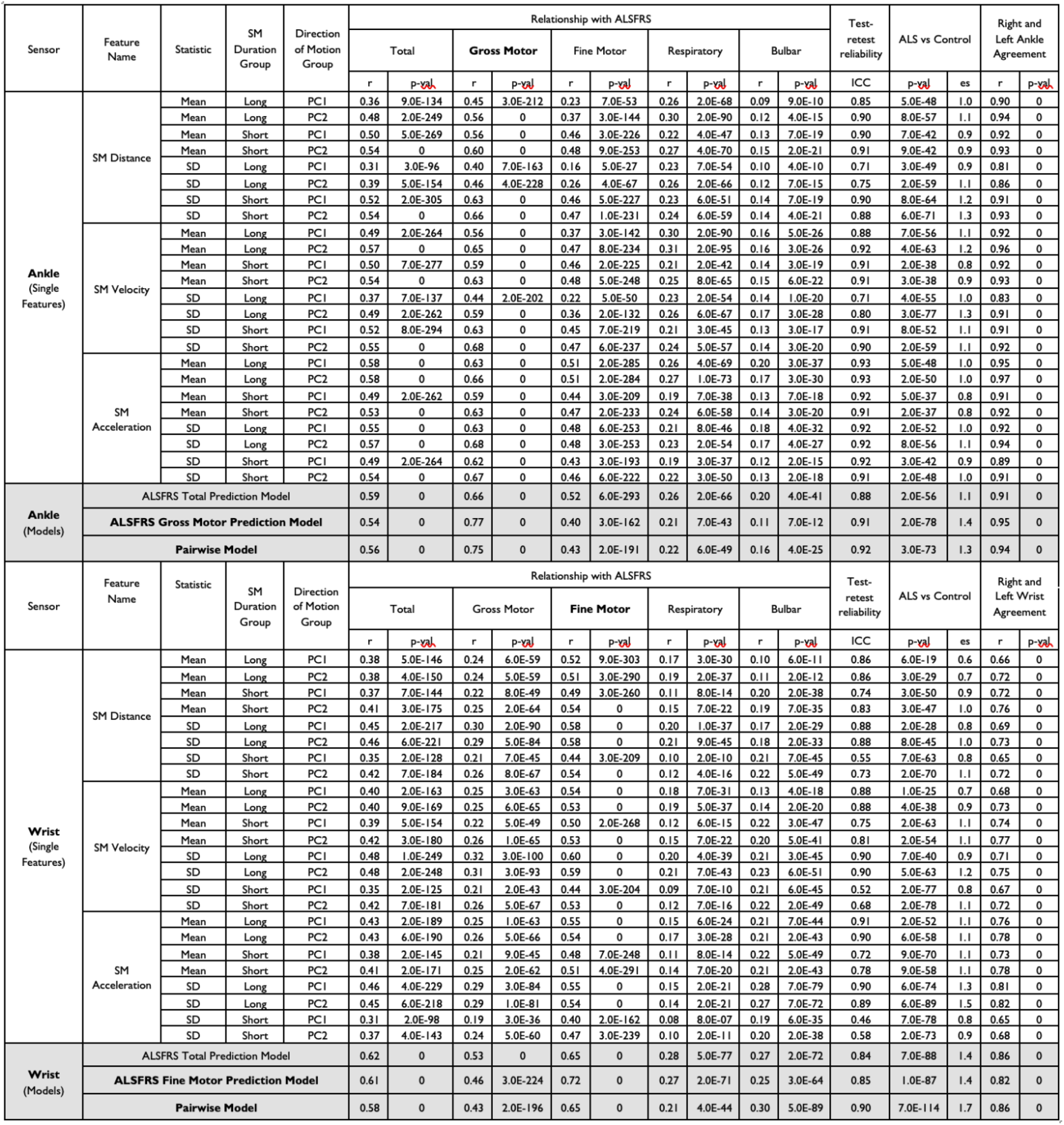
Cross-sectional properties of ankle and wrist submovement features and models. Abbreviations: SM – Submovement; SD – Standard deviation; AI – Activity Intensity; PC – Principal Component; ALSFRS-R – ALS Functional Rating Scale-Revised; ICC – Intraclass correlation coefficient; r – Pearson correlation coefficient; es – effect size.

**Figure 1.**
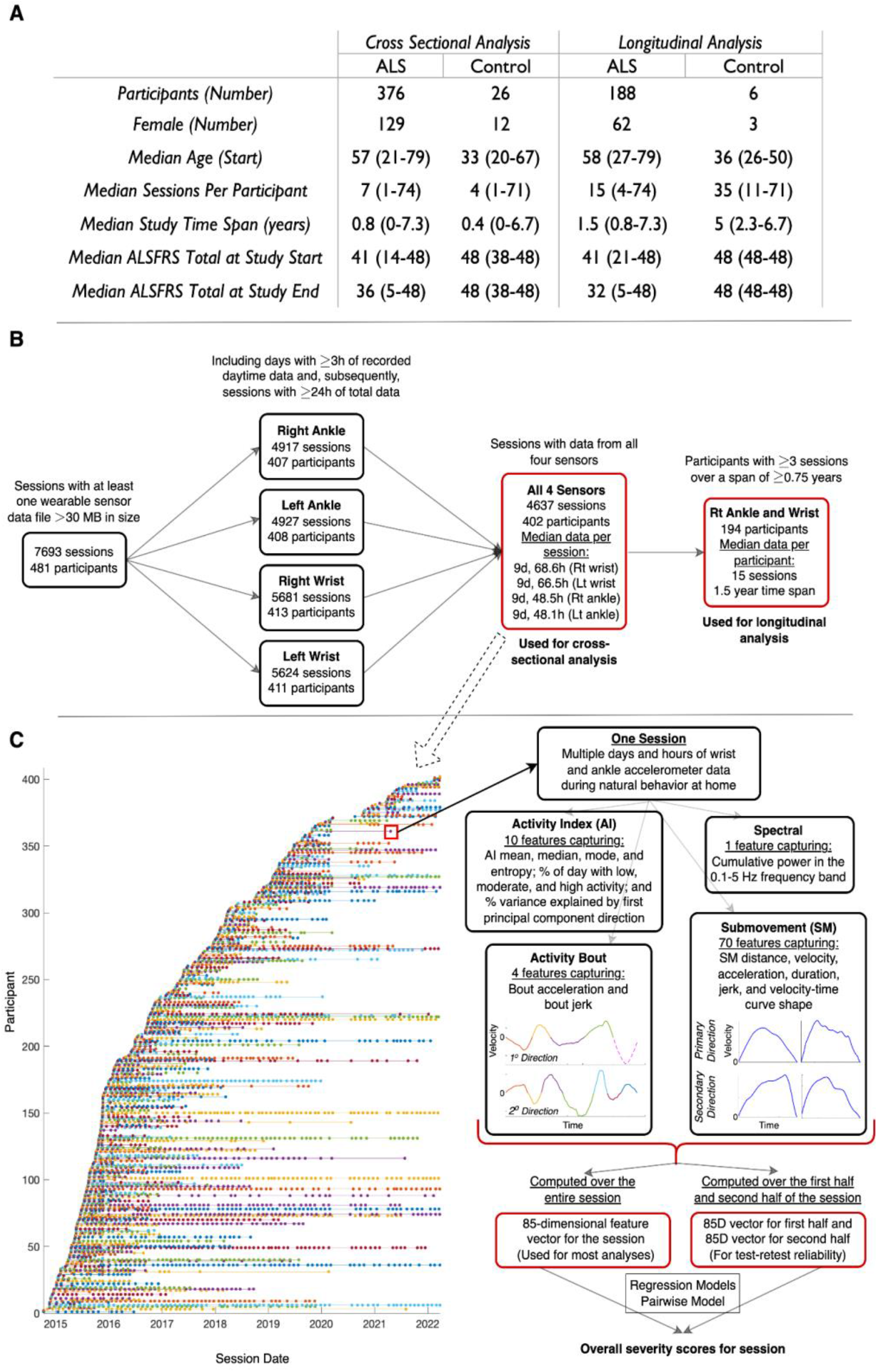
Overview of population and dataset. **A)** Participant clinical and demographic data with range of values provided in parentheses. **B)** Filtering steps for inclusion of sessions and participants used in cross-sectional and longitudinal analysis. **C)** Visualization of each participant’s session time points for data collection from 2014 to 2022, along with the movement features extracted from each session. Abbreviations: ALSFRS – ALS Functional Rating Scale-Revised; d – day; h – hour; Lt – left; Rt – right

### Overview of the pairwise comparisons severity estimation model

The task was to train a machine learning model that could combine information across the 85 movement features, previously shown to strongly reflect motor function in pediatric and adult ataxias^20,21^, to produce an ALS-specific composite measure that was sensitive to disease progression. The standard machine learning approach is to train a regression model to predict the clinical scale score (e.g., ALSFRS-R). However, the sensitivity of the model is then constrained by the sensitivity of the scale. In the “pairwise model” approach, the model is trained to learn the steepest direction of disease change in feature space based on longitudinal data, without using clinician or patient-reported information. This approach, described in Figure 2, can be applied to any disease that progresses over time. In addition to the pairwise model, linear regression models with L1-regularization^22^ were trained to predict ALSFRS-R total, ALSFRS-R gross motor subscore, and ALSFRS-R fine motor subscore, and were evaluated using five-fold cross-validation.

**Figure 2.**
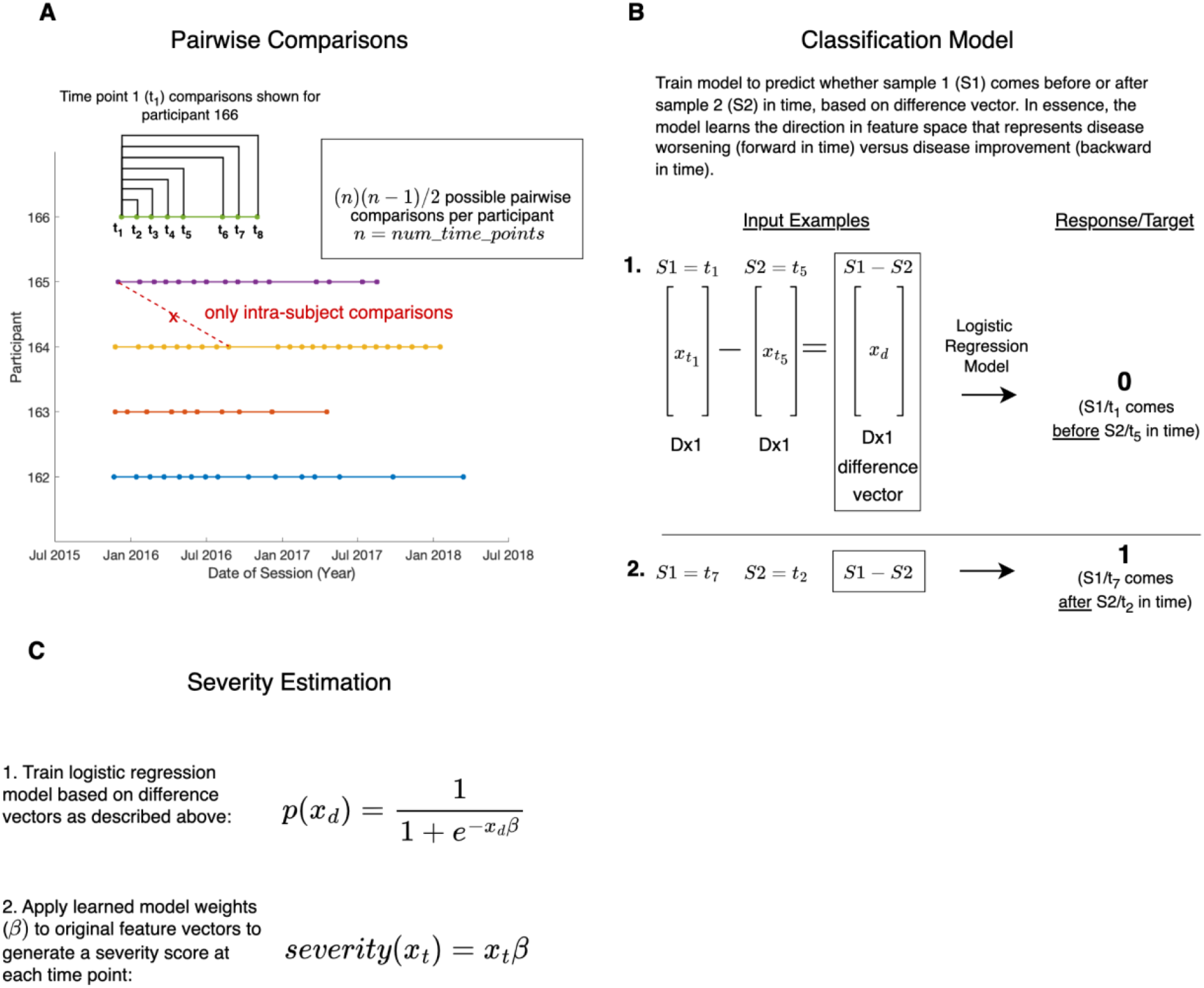
Overview of the pairwise comparisons model. **A)** Schematic showing the intra-subject pairwise session comparisons performed for participant 166’s first session. For each individual there are *n* ∗(*n* −*1*)/*2* possible pairwise comparisons (where *n* represents the number of time points or sessions for that individual). **B)** The model takes two 85-dimensional feature vectors or samples (S_1_ and S_2_) from a single individual as input, representing that individual’s motor function at two different points in time (t_m_ and t_n_). The element-wise difference between the two vectors is computed (S_1_-S_2_), representing the direction of change in feature space. This difference vector is the input to a binary classifier (logistic regression) which learns to predict whether the direction of change reflects disease progression (t_n_ is temporally after t_m_) or reflects disease improvement (t_n_ is temporally before t_m_). **C)** The learned logistic regression model parameters (representing the direction of disease progression) were then applied as linear weights to the original feature vectors to generate a score that reflects how far in the direction of disease progression the individual had traveled at that moment in time.

### Cross-sectional properties of ankle and wrist sensor data

Individual right ankle SM properties, including SM distance, velocity, and acceleration were significantly correlated with ALSFRS-R total (r = 0.31-0.58), demonstrated high test-retest reliability (ICC = 0.71-0.93), and were significantly different between ALS and control participants (effect size = 0.8-1.3, Table 1, top). All submovement properties were positively correlated with ALSFRS-R, indicating that submovement distances, peak velocities, and peak accelerations were smaller and less variable in individuals with more severe disease. Similarly, right *wrist* submovement (SM) properties were positively correlated with ALSFRS-R total (r = 0.31-0.48) and were significantly different between ALS and control participants (effect size = 0.6-1.5, Table 1, bottom). Long duration wrist submovements showed high test-retest reliability (ICC = 0.86-0.91), whereas short duration submovements had moderate test-retest reliability (ICC = 0.55-0.83). Ankle submovement properties were more strongly correlated with the ALSFRS-R gross motor subscore (r = 0.40-0.68) than with the ALSFRS-R fine motor subscore (r = 0.16-0.51), and were only weakly correlated with respiratory (r = 0.19-0.31) and bulbar (r = 0.09-0.20) subscores. Conversely, wrist submovement properties correlated more strongly with the ALSFRS-R fine motor subscore (r = 0.40-0.60) compared with ALSFRS-R gross motor subscore (r = 0.19-0.32), and also only weakly correlated with the respiratory (r = 0.08-0.21) and bulbar (r = 0.10-0.28) subscores. Both ankle and wrist submovements demonstrated good agreement between right and left limbs, however ankle right/left agreement (r = 0.81-0.97) was stronger than wrist right/left agreement (r = 0.65-0.82).

Machine learning models trained to learn a composite severity score based on right *ankle* movement features, correlated well with ALSFRS-R total (r = 0.54-0.59) and ALSFRS-R *gross* motor subscore (r = 0.66-0.77), had high test-retest reliability (ICC = 0.88-0.92), distinguished between ALS participants and controls (effect size = 1.1-1.4), and demonstrated strong right/left limb agreement (r = 0.91-0.95, see Table 1). For the right *wrist*, composite severity scores correlated well with ALSFRS-R total (r = 0.58-0.62) and ALSFRS-R *fine* motor subscore (r = 0.65-0.72), had high test-retest reliability (ICC = 0.84-0.90), distinguished between ALS participants and controls (effect size = 1.4-1.7), and demonstrated strong right/left limb agreement (r = 0.82-0.86, see Table 1). The ankle and wrist pairwise models had the highest test-retest reliability among the machine learning models (ICC = 0.92 and ICC = 0.90) and were the focus of longitudinal analysis. To understand which individual features were the most salient in the ankle and wrist pairwise models, we identified features that were in the top five (out of 85) in feature importance for all 5 cross-validation folds. For the right *ankle* pairwise model, the features included SM peak velocity (mean, PC2 direction, long duration SM group) and SM distance (mean, PC2 direction, long duration SMs). For the right *wrist* pairwise model the most salient features were SM peak velocity (mean, PC2 direction, *long* duration SMs) and SM peak velocity (mean, PC2 direction, *short* duration SMs).

### Longitudinal properties of ankle and wrist sensor data

The rate of change over time for each sensor-based composite score and ALSFRS-R score was modeled using linear regression, with the slope of the best fit line determining the rate of change^23^. To compare the rate of change of different scores, each with a different range of values, each score was z-scored and rate of change reported in z-score/standard deviations (SD) per year.

The rate of change of the pairwise model composite score was computed for each limb. Rate of change was highly consistent across right and left ankles (r = 0.87) and right and left wrists (r = 0.80, Figure 3A). There was lesser agreement (r = 0.52-0.56) between each upper and lower limb pair (e.g., right ankle versus right wrist). Individual-level trajectories demonstrated examples in which all four limbs progressed similarly over time (Figure 3B), the lower limb pair had similar trajectories but differed from the upper limbs (Figure 3C), and where the trajectory of one or two limbs deviated from the others (Figure 3D).

**Figure 3.**
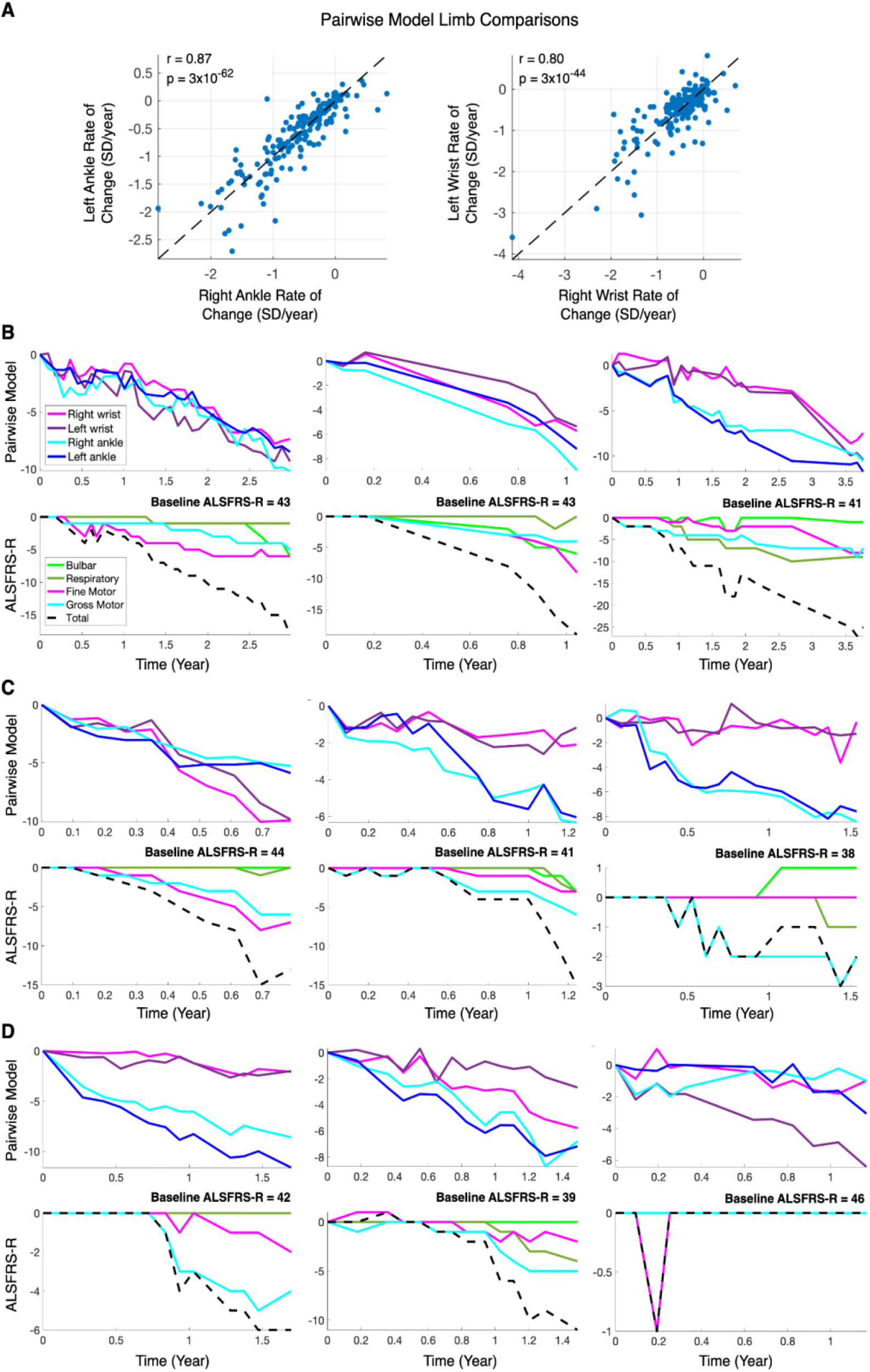
Longitudinal data from each limb. **A)** Agreement in rate of change of the pairwise model score between right and left ankles, and right and left wrists. Each point represents an individual with ALS. **B-D)** Longitudinal trajectories for nine individuals with ALS, with sensor-based pairwise model scores for each limb shown in the top panel and ALSFRS-R scores shown in the bottom panel. Individuals were observed to have similar trajectories for all four limbs **(B)**, similar trajectories for both ankles and both wrists **(C)**, or divergent trajectories for one or more limbs **(D)**. Abbreviations: SD – standard deviation; ALSFRS-R – ALS Functional Rating Scale-Revised

There was also congruence between lower limb pairwise model trajectories and ALSFRS-R gross motor subscore trajectories and between upper limb and ALSFRS-R fine motor subscore trajectories (Figure 3B-D). The population-level agreement between the *right ankle* pairwise model rate of change and ALSFRS-R *gross motor* rate of change (r = 0.73, p = 1.5×10^−33^) was stronger than the agreement with ALSFRS-R fine motor (r = 0.56, p = 1.4×10^−17^), and the *right wrist* pairwise model rate of change showed stronger agreement with ALSFRS-R *fine motor* (r = 0.73, p = 1.1×10^−33^) compared to ALSFRS-R gross motor rate of change (r = 0.60, p = 4.2×10^−20^).

Next, for each participant, the pairwise model rate of change was combined over the four limbs by either taking the average rate of change or the maximum rate of change. When taking the *average* of the four limbs, the pairwise model rate of change had strong agreement with ALSFRS-R total rate of change (r = 0.71), gross motor subscore rate of change (r = 0.75), and fine motor subscore rate of change (r = 0.68, Figure 4A), and weak agreement with respiratory and bulbar subscores (r = 0.38 and r = 0.45, respectively). Similarly, when taking the limb with the *maximum* rate of change, the pairwise model had strong agreement with ALSFRS-R (r = 0.69), gross motor subscore (r = 0.75), and fine motor subscore (r = 0.69, Figure 4B), and weak agreement with respiratory and bulbar subscores (r = 0.34 and r = 0.43, respectively). The sensor-based pairwise model, which was trained to estimate disease severity without knowledge of ALSFRS-R scores, had strong rate-of-change agreement with the regression model trained to estimate ALSFRS-R total score, regardless of whether the average of the four limbs or the limb with the fastest progression rate was used (r = 0.92 for both, Figure 4A,B). Thus averaging or taking the maximum rate of change across the four limbs produced equally robust and consistent measures of disease progression.

**Figure 4.**
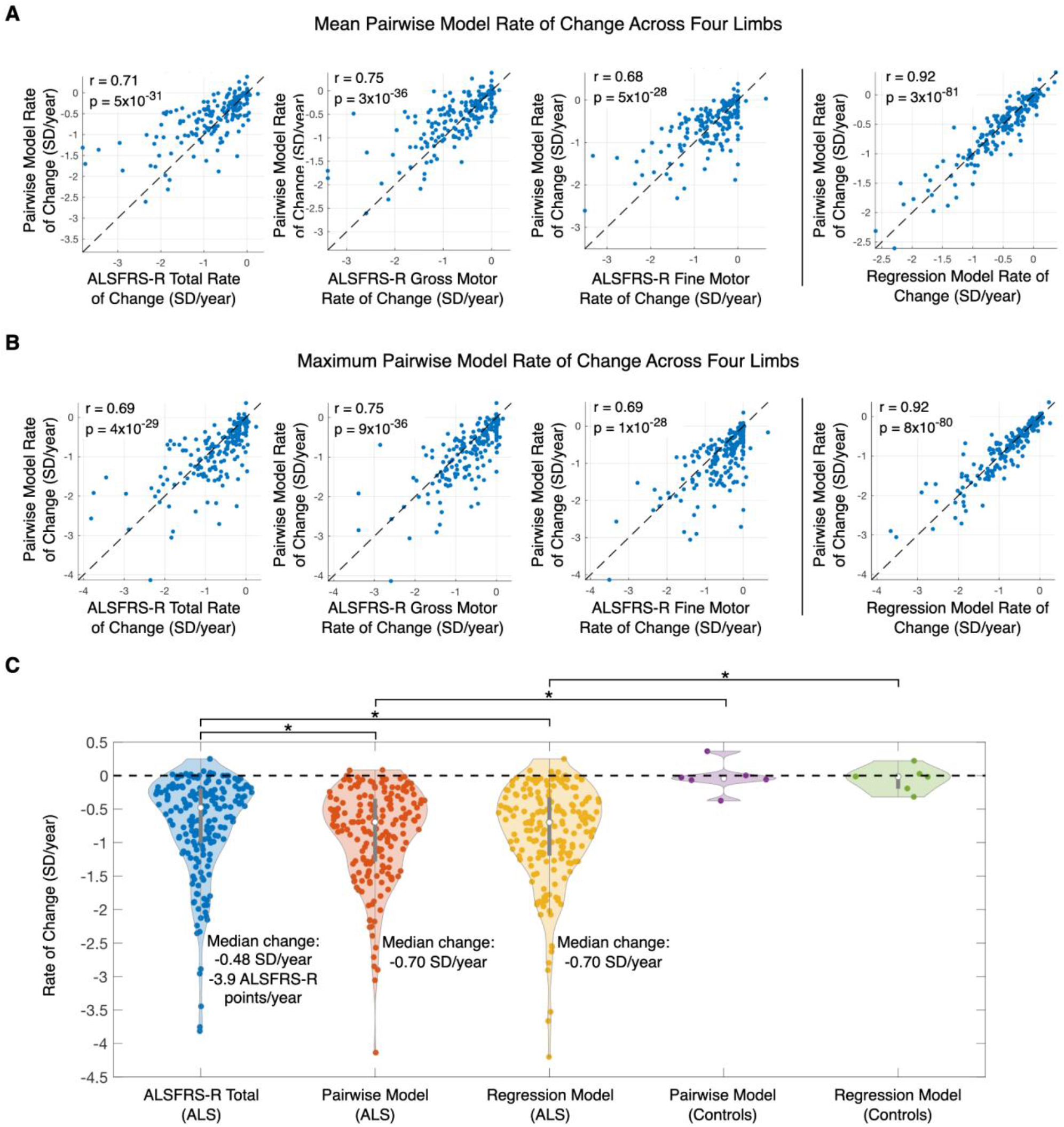
Rate of change comparison between sensor-based models and ALSFRS-R. **A)** Mean pairwise model rate of change across four limbs compared with ALSFRS-R total score, gross motor subscore, and fine motor subscore (left) and compared with the mean regression model rate of change (right). **B)** Maximum pairwise model rate of change across four limbs compared with ALSFRS-R total score, gross motor subscore, and fine motor subscore (left) and compared with the maximum regression model rate of change (right). **C)** Violin plot comparing the distributions of ALSFRS-R rate of change, pairwise model rate of change (using fastest progressing limb), and regression model rate of change (using fastest progressing limb). For **A-C**, each point represents a participant. Abbreviations: SD – standard deviation; ALSFRS-R – ALS Functional Rating Scale-Revised

When taking the maximum rate of change, points shift downward with respect to the y=x line (Figure 4A versus 4B) indicating increased sensitivity of the sensor-based model to disease change in comparison with ALSFRS-R total. Using the maximum rate of change, the pairwise model and the regression model progressed faster over time (−0.70 SD/year for both) than ALSFRS-total (−0.48 SD/year, p = 0.007 and p = 0.017, respectively; Figure 4C). Pairwise model and the regression model scores did not progress for control participants and were significantly different between ALS and control participants (p = 0.0004 and p = 0.0006, respectively; Figure 4C). When using the mean rate of change of all four limbs, the pairwise model and ALSFRS-R total score were equally sensitive (−0.45 SD/year versus -0.48 SD/year, p = 0.12) and ALSFRS-R total score was slightly more sensitive than the regression model (−0.48 SD/year versus -0.44 SD/year, p = 0.037).

## Discussion

We have shown that data from inexpensive sensors worn on limbs at home during natural behavior can produce reliable, sensitive, and interpretable measures of gross and fine motor function in individuals with ALS. Ankle movement properties derived from accelerometer data were highly consistent across right and left ankles and were in agreement with gross motor function as assessed on ALSFRS-R, both in terms of cross-sectional severity and in terms of rate of change over time. Similarly, wrist movement properties were highly consistent across right and left wrists and were in agreement with fine motor function on ALSFRS-R. Although there was strong right-left limb agreement at a population level, arm-leg agreement showed only moderate agreement, and some individuals were observed to have different rates of progression for each limb. Taking the score of the limb with the maximum progression rate produced a motor outcome measure that was consistent with but more sensitive than the current primary outcome measure in most ALS trials (ALSFRS-R).

The analysis approach for quantifying motor function in ALS centered on extraction and characterization of motor primitives called submovements during *natural behavior*, which was previously developed for quantifying motor function in ataxia-telangiectasia^20^ and adult cerebellar ataxias^21^. There is evidence that motor control is achieved by combining submovements to compose complex voluntary motor behaviors^24–27^ and that submovements change in a consistent manner with the state of the motor system. With motor development, infants’ reaching trajectories become straighter, and submovements decrease in number and increase in duration^28^. Older individuals compensate for greater noise and lower perceptual efficiency by increasing the number of submovements and decreasing the velocity of submovements during accuracy-constrained movement tasks^29^. During recovery from stroke, the number of submovements decreases and their temporal overlap increases giving rise to smoother trajectories during point-to-point movements^30^. In individuals with ataxia, wrist submovements during a point-to-point reaching task^31^ and ankle submovements during a gait task^32^ become smaller and slower with increasing ataxia severity. Similarly, during natural at-home behavior, ankle submovement distance, peak velocity, and peak acceleration are smaller in adults with spinocerebellar ataxias and multiple system atrophy compared to controls and become progressively smaller and less variable as self-reported function decreased and ataxia severity increase^21^. The submovement analysis approach contrasts with a prior analysis of task-free, at-home measurement in 42 individuals with ALS using waist-worn accelerometers, which quantified overall activity levels (e.g., activity count, percent of day active)^19^. Although overall motor activity is a pertinent outcome in ALS, it is reliant on full day sensor wear and is likely more susceptible to day-to-day changes in behavioral context (e.g., travel, systemic illness, sleep quality), requiring careful consideration of reliability.

Based on our literature review, limb submovement properties have not been previously studied in ALS. Several studies, however, have investigated the relationship between muscle strength (a direct cause of motor impairment in ALS^33^) and submovement characteristics. In a heterogeneous population of individuals with motor impairments (e.g., spinal cord injury, cerebral palsy, stroke), participants were asked to perform a computer-based pointing task and a mechanical dynamometer was used to measure grip strength and pinch strength^34^. The authors found that the number of submovements per pointing movement was negatively correlated with grip strength (the movement was composed of smaller submovements as grip strength decreased) and that the velocity of movement was directly proportional to grip strength^34^. In another study of individuals with hemiparesis secondary to stroke, it was found that peak arm reaching velocity was influenced most by shoulder, elbow, and wrist flexor and extensor muscle strength (58% of variance explained), measured using a hand-held dynamometer^35^. In a study of individuals without motor disability, submovement organization was examined as participants tracked a small or large dot on a screen with a pen placed on a digitizer tablet, while simultaneously recording activity from muscles in the neck and upper extremity using surface electrodes^36^. When tracking the smaller target, extensor and flexor muscles of the forearm activated more strongly, and submovements were found to have increased peak velocities^36^.

These studies support that there is a robust relationship between muscle strength and submovement properties, in particular peak velocity. Consistent with these studies, we found that wrist and ankle submovements from individuals with ALS had smaller velocities, accelerations, and distances traversed. Submovement peak velocity was the only highly selected feature in both the right ankle and the right wrist pairwise models, demonstrating its importance for measuring disease progression ALS. This supports a model in which muscle weakness and decreased muscle activation caused by motor neuron pathology gives rise to slower and smaller submovements during everyday limb movement. Further supporting this model, are the parallels in left-right symmetry observed in the present study with the left-right symmetry observed in large studies of hand-held dynamometry (HHD)^23^ and Accurate Test of Limb Isometric Strength (ATLIS)^37^ in ALS. Individual arm and leg muscles were found to correlate strongly with the identical muscles on the contralateral side, both in terms of cross-sectional strength measurements (r = 0.65 to 0.90) and also in terms of rate of change over time (r = 0.43 to 0.82)^23^. We observed similar side-to-side cross-sectional and rate of change symmetry in individual submovement features (cross-sectional r = 0.65 to 0.97) and composite models (cross-sectional r = 0.82 to 0.95; rate of change r = 0.80 to 0.87). Interestingly, side-to-side cross-sectional symmetry of the leg was stronger than side-to-side symmetry of the arm here and in the HHD study. This may have implications for how ALS disease pathology spreads and highlights a potential future application of this technology in characterizing phenotypic spread across limbs in a continuous and granular fashion, for example in presymptomatic gene carriers. This also supports that submovement characteristics may be a suitable proxy for muscle strength in ALS, and offers an advantage over HHD and ATLIS of being able to measure strength continuously over multiple days, during the individual’s own selection of behaviors, and without relying on participant effort or evaluator training and strength. Thus, it may produce more reliable, ecologically valid, and scalable measures of muscle strength and motor function. It may also apply to other neurological conditions that affect muscle strength. A future study that collects HHD and/or ATLIS measurements along with submovements from accelerometer data would help clarify the relationship between strength and submovements in ALS.

As discussed above, strong side-to-side correlations of ankle and wrist submovement features and composite models were observed. This is consistent with previously reported strength measurements in ALS^23,37^, but also highlights the robustness of the submovement measures that are generated independently from each limb’s movement during natural behavior at home. Ankle submovement measures correlated strongly with ALSFRS-R gross motor subscore (both cross-sectional scores and rate of change) and wrist submovement measures correlated strongly with ALSFRS-R fine motor subscores. We found high test-retest reliability of the sensor-based features and composite models. Finally, two machine learning models trained based on different information (pairwise model trained on longitudinal change; regression model trained on ALSFRS-R) generated composite scores that had strong agreement in rate of disease progression (r = 0.92). These properties support that sensor-derived submovements obtained during natural behavior provide highly robust measures of disease severity for each limb. Since each limb can be reliably and independently measured, these data support the use of the fastest progressing limb’s rate of progression in order to obtain a personalized overall measure that is more sensitive for measuring disease change than ALSFRS-R and which may be more responsive to therapeutic intervention. However, the choice of if and how to combine severity measures from each limb can be determined based on the clinical application as well as on the individual’s prior clinical trajectory.

Two different supervised machine learning approaches were used to create composite measures of overall motor impairment for each limb based on the collection of sensor-based movement features. One used the traditional approach of training a regression model to predict severity as measured by ALSFRS-R. The other approach learned the trajectory of disease progression (in feature space) from the longitudinal data and computed how far the individual had moved along that trajectory without ever having access to rating scale data (i.e., pairwise model). Despite the very different training approaches, both models were highly consistent in their estimates of progression rate (r = 0.92) and were similarly consistent with ALSFRS-R total’s progression rate (r = 0.69 and r = 0.71). The pairwise model was highlighted in analysis for three main reasons: 1) it had higher reliability than the regression models, 2) the consistency with ALSFRS-R in cross-section and in rate of change was striking given that it had no chance to “overfit” to the clinical score, and 3) the pairwise modeling approach may be useful for other diseases where the existing clinical rating scale is less sensitive for capturing disease change. Furthermore, the pairwise modeling approach can be extended in a number of ways, for example by filtering comparisons, changing the type of classifier used, and aggregating data across multiple disease populations.

The large and longitudinal dataset generated by the ALS-TDI Precision Medicine Program, consisting of 376 individuals with ALS who wore four sensors for multiple hours and days at home and with 188 participants who wore the four sensors longitudinally over a minimum of 0.75 years (median of 15 times over 1.5 years), supports the feasibility of the at-home passive data collection approach from both a patient and clinical operations perspective.

There were some limitations to the study. There was a relatively small number of controls included in the study and the controls were not age matched. However, the size and characteristics of the control sample do not affect the main conclusions of the study. There was heterogeneity in the number of hours each participant wore the sensors at home. This was mitigated in part by only including days in which sensors were worn for at least 3 hours. We anticipate higher reliability estimates of all sensor-based measures if participants are explicitly asked to wear the sensor throughout the entire day with exception of bathing (and night if possible). Finally, as expected, the severity estimates based on limb movement did not correlate well with bulbar and respiratory function. These functions are represented in ALSFRS-R and other digital strategies (e.g., video-based analysis of facial movement or speech analysis^17,38–40^) are needed to quantify these important motor domains in ALS.

In summary, we have shown that a submovement-based analysis of natural behavior at-home using wearable sensors produces interpretable, reliable, sensitive, and ecologically valid measures of gross and fine motor function in ALS. This technology has properties that support its use as a novel outcome measure in ALS clinical trials with the potential to reduce the cost and size of future trials. The use of inexpensive sensors, worn at home with minimal instruction and no eligibility limitations, could increase access to clinical trials and support virtual clinical trials in ALS. It may also support the routine clinical care of individuals with ALS by providing clinicians and patients with an objective and reliable motor assessment that can be passively obtained at home with relatively low burden and cost.

## Methods

This research study was conducted in accordance with the ethical principles posited in the Declaration of Helsinki - Ethical Principles for Medical Research Involving Human Subjects. Protocol approval was provided by the institutional review board (ADVARRA CIRBI). Every participant consented to participate in this research by signing an IRB approved informed consent form.

### Wearable Sensor Data Processing and Feature Types

Continuous triaxial accelerometer data collected at 30 Hz was obtained from Actigraph GT3X devices (one for each limb). In prior work, each participant’s wearable sensor data were manually partitioned into day and night segments based on changes in each participant’s daily activity level represented in the accelerometer data^20,21,41^. However, given the large size of this dataset, day segments were automatically partitioned to include data collected between 7:21 am and 11:27 pm^42^, while accounting for each individual’s time zone. Data analysis focused on daytime segments. Gravity and high frequency noise were removed from the acceleration time-series using a sixth order Butterworth filter with cutoff frequencies of 0.1 and 20 Hz.^20,21,41,43^

Several classes of features were extracted from daytime ankle and wrist sensor data as in prior work^20,21^. These included *total power* in the 0.1-5 Hz frequency range and features based on the distribution of *activity intensity* computed in 1-second time bins. Features were also extracted from “activity bouts’’ and from submovements. Supplementary Table 1 provides a description of the 85 features extracted from ankle and wrist sensor data. Based on prior work, single feature analysis was performed on a subset of 24 submovement features of interest as described in the main text.

### Severity Estimation Models

Supervised machine learning approaches were used to create composite severity scores that aggregate over the 85 movement features. Separate models were trained for each limb. The pairwise comparison approach is described in Figure 2 and the main text. To ensure that the pairwise model did not inadvertently learn longitudinal changes resulting from changes in device settings, comparisons were only allowed between sessions that had the same critical firmware version (where raw data were collected in an identical way). Five-fold cross-validation was used: for each fold comparisons from 80% of ALS participants were used to train a classification model and the model weights were applied to data from the held-out 20% of participants to generate severity scores for each session. Additionally, we trained linear regression models with L1 regularization (i.e., lasso regression)^22^ to predict ALSFRS-R total, ALSFRS-R gross motor subscore (ankle sensor data only), and ALSFRS-R fine motor subscore (wrist sensor data only). Five-fold cross-validation was also used to evaluate performance of the regression models. For both the pairwise models and the regression models, each feature was z-score transformed prior to model training such that feature value ranges and model weights were comparable. Pearson correlation coefficient was used to measure performance, with each model compared with ALSFRS-R.

### Statistical Analyses

Statistical analyses were completed in MATLAB (Mathworks, Natick, MA) and SPSS (IBM Corp., Armonk, NY). The Mann-Whitney *U*-test was used to determine individual feature differences between disease and control groups and Cohen’s *d* was used to measure effect size. The Mann-Whitney *U*-test was also used to determine differences in rate of change between different assessments. The Benjamini-Hochberg method was used to adjust for multiple comparisons and corrected p-values are reported^44^. Corrected p-values < 0.05 were considered significant. Single measure intraclass correlation coefficients (ICCs) were used to determine the test-retest reliability of features and composite scores. To evaluate reliability of sensor-based features, features were computed from data recorded in first half of the days in the session and the second half of the days in the session, separately, and ICCs were computed using a 2-way mixed effects model.^45^ Pearson correlation coefficients and p-values were used to evaluate the relationship between sensor-based features and ALSFRS-R. As above, the Benjamini-Hochberg method was used to adjust for multiple comparisons^44^.

## Data Availability

Data included in this study will be shared by request from any qualified investigator.

## Data availability

Data included in this study will be shared by request from any qualified investigator.

## Acknowledgements

The authors thank James Berry and Katherine Burke for helpful discussions. We also thank the community of people with ALS who contributed data to these studies.

## Author Contributions

F.V. conceived of the translational research program that resulted in the data collected and analyzed in this manuscript. A.S.G., S.P., A.P., and F.V. conceived of the study objectives. A.P. and F.V. contributed to data collection efforts. S.P. performed ingestion and preprocessing of the dataset. A.S.G. performed analysis of the dataset. A.S.G., S.P., A.P., and F.V. contributed to the interpretation of the results. A.S.G. took the lead in writing the manuscript. All authors provided critical feedback and helped shape the research, analysis, and manuscript.

